# Insights into smouldering MS brain pathology with multimodal diffusion tensor and PET imaging

**DOI:** 10.1101/2020.01.09.20017012

**Authors:** Svetlana Bezukladova, Jouni Tuisku, Markus Matilainen, Anna Vuorimaa, Marjo Nylund, Sarah Smith, Marcus Sucksdorff, Mehrbod Mohammadian, Virva Saunavaara, Sini Laaksonen, Johanna Rokka, Juha O. Rinne, Eero Rissanen, Laura Airas

## Abstract

**Objective:** To evaluate *in vivo* the co-occurrence of microglial activation and microstructural white matter damage in multiple sclerosis (MS) brain, and to examine their association with clinical disability.

**Methods:** 18-kDa translocator protein (TSPO) brain PET imaging was performed for evaluation of microglial activation by using the radioligand [^11^C](R)-PK11195. TSPO-binding was evaluated as the distribution volume ratio (DVR) from dynamic PET images. Diffusion tensor imaging (DTI) and conventional MRI were performed at the same time. Mean fractional anisotropy (FA) and mean (MD), axial (AD) and radial (RD) diffusivities were calculated within the whole normal appearing white matter (NAWM) and segmented NAWM regions appearing normal in conventional MRI. 55 MS patients and 15 healthy controls were examined.

**Results:** Microstructural damage was observed in the NAWM of MS brain. DTI parameters of MS patients were significantly altered in the NAWM, when compared to an age- and sex-matched healthy control group: mean FA was decreased, and MD, AD and RD were increased. These structural abnormalities correlated with increased TSPO binding in the whole NAWM and in the temporal NAWM (*p*<0.05 for all correlations; *p*<0.01 for RD in the temporal NAWM). Both compromised WM integrity and increased microglial activation in the NAWM correlated significantly with higher clinical disability measured with expanded disability status scale (EDSS).

**Conclusions:** Widespread structural disruption in the NAWM is linked to neuroinflammation, and both phenomena associate with clinical disability. Multimodal PET and DTI imaging allows *in vivo* evaluation of widespread MS pathology not visible using conventional MRI.

Supplemental data: Table e-1. Clinical and conventional MRI parameters of healthy controls and multiple sclerosis patients

Table e-2. The effect of number of gradients (64 vs. 33) on DTI-derived measures

Table e-3. Associations of DTI and TSPO-PET measures in the segmented NAWM with disability and disease severity among MS patients (n=55).

## INTRODUCTION

Demyelination, inflammation, and neurodegeneration of brain, spinal cord and optic nerves are the hallmarks of multiple sclerosis (MS) pathogenesis.^1^ Conventional magnetic resonance imaging (cMRI) is used for evaluation of focal inflammatory activity and diffuse atrophy for diagnostics and follow-up of MS.^2^ Even though cMRI has a well-established role in clinical setting as well as in the evaluation of treatment effect in longitudinal studies with disease modifying therapies (DMT), this imaging modality alone is not able to provide the full picture on MS pathology *in vivo*. In order to enhance the detection of diffuse pathological alterations associated with advanced MS, several methodologies have been applied. With the implementation of diffusion tensor imaging (DTI), it is possible to estimate focal diffusivities and diffusion anisotropy, thereby revealing brain parenchymal alterations at the microstructural level, undetectable in cMRI.^3^ *In vivo* molecular imaging of neuroinflammation using positron emission tomography (PET) with translocator protein (TSPO) binding radioligands provides on the other hand quantifiable molecular data on the activated microglia in the brain.^4^ The primary objective of this study was to evaluate the diffuse pathology in the NAWM using DTI with simultaneous evaluation of microglial activation using TSPO-PET in a large cohort of MS patients. We propose that the combination of these imaging modalities will enable better understanding of the “hidden” MS pathology not visible using cMRI.

## METHODS

### Study Protocol Approval

The study protocol has been approved by the Ethics Committee of the Hospital District of Southwest Finland. Informed consent was obtained from all participants before entering the study according to the principles of the Declaration of Helsinki.

### Participants

In total, a cohort of 55 MS patients (40 relapsing remitting MS (RRMS), 15 secondary progressive MS (SPMS), age 28–64 years, expanded disability status scale (EDSS) 1–6.5) was examined. 20 RRMS and 15 SPMS patients had no immunomodulatory treatment at the time of imaging, while other 20 RRMS patients were using DMTs during imaging (3 patients were taking dimethyl fumarate, 5 patients were on fingolimod treatment, 6 patients had interferon beta 1-a treatment, 4 patients were receiving glatiramer acetate and 2 were on teriflunomide). All patients with DMT were clinically stable and free from relapses at the time of imaging. Data from fifteen healthy controls (age 21-58 years) were included for comparison. The healthy controls were imaged with [^11^C](R)-PK11195 and DTI using similar protocols except for five subjects, from whom only conventional MRI and DTI data were available. The demographics and clinical characteristics of the study subjects are shown in Table e-1.

Clinical evaluation and neurological status including the evaluation of disability with EDSS and disease severity with multiple sclerosis severity scale (MSSS) was performed for all MS patients.

### Data acquisition

Magnetic resonance imaging was performed with 3T MRI Phillips Ingenuity (Philips Healthcare, Cleveland, OH) scanner. The MRI protocol included T1, T2-weighted, FLAIR and DTI sequences. For DTI sequences, 33 gradient directions were utilized for imaging of 20 MS patients (15 SPMS, 5 RRMS), while DTI with 64 gradient directions was applied in the rest. All other DTI parameters were equal with b-value=1000s/mm^-2^, TR/TE=9500/120 ms, FOV=256×256 mm, spatial resolution 2×2×2 mm, acquisition matrix 128×128 mm, flip angle=90° and acceleration factor 2.

Dynamic 60 minutes [^11^C](R)-PK11195-PET imaging was performed with ECAT HRRT scanner (CTI, Siemens Medical Solutions, Knoxville, TN). The radioligand was administered as a smooth, intravenous bolus injection, the target dose being 500 MBq. The mean (SD) of injected activity was 473.6 (59.5) MBq for MS patients and 498.8 (7.9) MBq for control subjects, with no significant differences between groups.

### Data pre-processing and analysis

The T2 hyperintense lesions were first identified from the FLAIR images using Lesion Segmentation Toolbox (version 2.0.15).^5^ The T1 image was filled with the manually corrected lesion masks, following a region of interest (ROI) delineation with Freesurfer 6.0 software. To create a NAWM ROI, cerebellar white matter and T2 hyperintense lesions were excluded from white matter ROI. Next, six subregions of NAWM (deep, cingulate, frontal, temporal, occipital and parietal NAWM) were derived from Freesurfer WM parcellation.^6^ DWI data were pre-processed with ExploreDTI for motion, eddy current and EPI/susceptibility induced distortion correction.^7^ Diffusion tensor estimation method was set to RESTORE (robust estimation of tensors by outlier rejection) approach.^8^ After DTI data pre-processing, four maps of interest (FA, MD, AD, RD) were reconstructed from the diffusion tensor map and co-registered in SPM12 (The Wellcome Centre for Human Neuroimaging, University College London) running in MATLAB (The Mathworks, Natick, MA) to corresponding T1-weighted image. Finally, all images were spatially normalised into the MNI152 space.

Due to pooling of DTI data with different number of gradients (33 vs. 64), we performed a test whether the number of gradients have an impact on DTI scalar indices. We reduced the number of gradients from 64 to 33, using ExploreDTI function ‘Shuffle/select 3D volume(s) in 4D *.nii file(s)’ and re-evaluated DTI scalars with lower number of gradients for 5 patients. Since DTI contrast is mainly dependent on the diffusion weighting factor (b-value) and echo time (TE)^9^, DTI-derived indices for both gradients schemes were comparable (Supplementary material, Table e-2.). None of the pairwise comparisons reached a statistically significant difference when performing a Student’s t-test between DTI-derived scalars obtained with 64 and 33 gradients.

PET images were reconstructed using 17-time frames (total of 3600 seconds) as described previously.^10^ Thereafter, pre-processing was carried out in similar manner as reported earlier.^11^ The dynamic PET images were co-registered to 3D T1 MR images, and then all data were resliced to 1mm voxel size. For evaluating [^11^C](R)-PK11195 binding, time-activity curves (TACs) from the seven ROIs were extracted from the pre-processed PET images using the same NAWM masks as in the DTI analyses including the global NAWM and the six NAWM subregions.

Regional TSPO radioligand binding was evaluated as distribution volume ratio (DVR) by using a reference region extracted with the supervised cluster algorithm (SuperPK software) as described earlier.^10^ The reference tissue–input Logan method, within 20-60 minutes time interval, was applied to the regional TACs using the clustered gray matter reference tissue input. Additionally, the modelling was performed at voxel level, where the parametric binding potential (BP_ND_) maps were calculated using a basis function implementation of simplified reference tissue model^12^ with 250 basis functions. The resulting parametric maps were further transformed to DVR (DVR=BP_ND_ +1) and normalized into MNI152 space in SPM12.

### Statistical methods

The statistical analyses of DTI, PET imaging and clinical parameters were performed with R software (version 3.5.2). The normality distribution of the data was evaluated with the Shapiro-Wilk test. The nonparametric Mann–Whitney U test was chosen for the evaluation of the group differences of non-normally distributed data and in groups with a low number of subjects. Holm method was used to adjust the *p*-values in case of the multiple comparisons. Student’s t-test was chosen to test for age difference between groups. PET data were analysed with analysis of covariance (ANCOVA) with age as covariate, and with Tukey’s HSD test for the adjustment for the multiple comparisons due to significantly different age between subgroups (mean (SD) age for HC, RRMS and SPMS: 37.2 (10.9), 45.6 (6.80) and 50.4 (9.23), respectively; *p*=0.019 for SPMS vs. HC and *p*=0.013 for RRMS vs. HC). The correlational analyses between variables of interest were analysed with Spearman’s nonparametric correlation test, where *p*-values adjusted using the Holm method for the number of ROI’s (6) in brain, separately for each of the DTI parameters or DVR values, and for EDSS and MSSS. Correlational analyses were not adjusted for age, as diffusion parameters and DVR values did not show a significant correlation with age (data not shown).

### Voxel-wise image analysis of the MS patient data

Pearson’s correlation coefficients were calculated at voxel level in the NAWM (excluding the lesions) and in the whole WM (including the lesions) between normalized FA and DVR images of 54 MS patients. One subject was excluded from analysis due to DTI image data artefacts. Before the correlation analysis, the parametric images were smoothed with Gaussian 8mm FWHM (full width at half maximum) filter to compensate for anatomical variability in image normalization and to improve signal-to-noise ratio. The resulting *p* values were corrected for multiple comparisons using false discovery rate (FDR) with significance level *p*<0.05.

### Data Availability Statement

The raw data used in the preparation of this article can be shared in anonymized format by request of a qualified investigator.

## RESULTS

### Increased microglial activation in the NAWM of MS brain

SPMS patients had higher [^11^C](R)-PK11195 DVR values as an indication of increased microglial activation in the whole NAWM compared to healthy controls and to RRMS patients [1.262 (0.046) vs. 1.194 (0.026); *p*=0.0002 and vs. 1.222 (0.039); *p*=0.003, respectively]. Within the NAWM subregions, the [^11^C](R)-PK11195 DVR was higher in SPMS patients compared to healthy controls and to RRMS patients in frontal NAWM [1.328 (0.081) vs. 1.199 (0.033); *p*<0.0001 and vs. 1.245 (0.046); *p<*0.0001, respectively] and was significantly higher in RRMS compared to HC; *p*=0.048. [^11^C](R)-PK11195 DVR was also increased in SPMS vs. HC and vs. RRMS in cingulate NAWM [1.204 (0.051) vs. 1.117 (0.047); *p*=0.0004 and vs. 1.141 (0.054); *p*=0.0006, respectively] and in deep NAWM [1.207 (0.077) vs. 1.125 (0.034); *p=*0.004 and vs. 1.151 (0.058); *p=*0.009, respectively]

### Structural white matter changes in MS patients

In the DTI analyses, we observed a trend for lower mean FA close to statistical significance in the whole NAWM among the MS patients compared to healthy controls [mean (SD) 0.341 (0.026) vs. 0.356 (0.017*); p*=0.064] (Figure 2A). In the diffusivity indices of the whole NAWM, no significant differences were observed.

**Figure 1.**
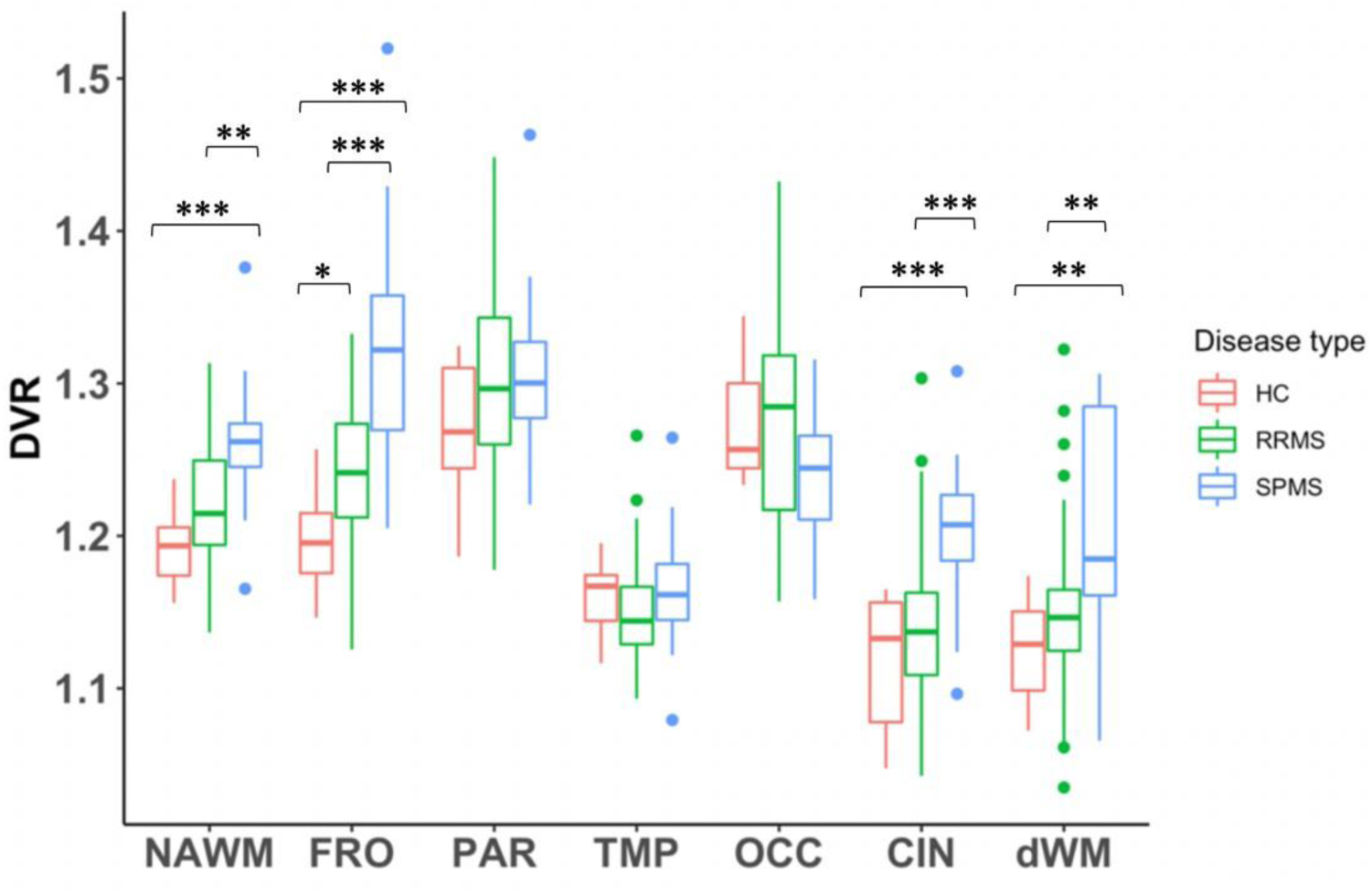
[^11^C](R)-PK11195 binding in the global and segmented NAWM in MS patients compared to healthy controls. ROI-specific binding of [^11^C](R)-PK11195 measured as DVR in the global NAWM and in segmented NAWM of 55 MS patients, compared to 10 HC. Boxplots show median DVR values with first and third interquartile. Data beyond end of whiskers are outliers and plotted as points. The pairwise comparisons were performed with the analysis of covariance (ANCOVA) with age as covariate, followed by Tukey’s post-hoc adjustment for multiple comparisons, and were considered statistically significant at the level of *p*<0.05. CIN=cingulate; DVR = distribution volume ratio; dWM=deep normal appearing white matter; FRO=frontal; NAWM=global normal appearing white matter; OCC=occipital; PAR=parietal; RRMS=relapsing remitting multiple sclerosis; SPMS=secondary progressive multiple sclerosis; TMP=temporal; *\* p*<0.05; ** *p*<0.01; *** *p*<0.001.

**Figure 2.**
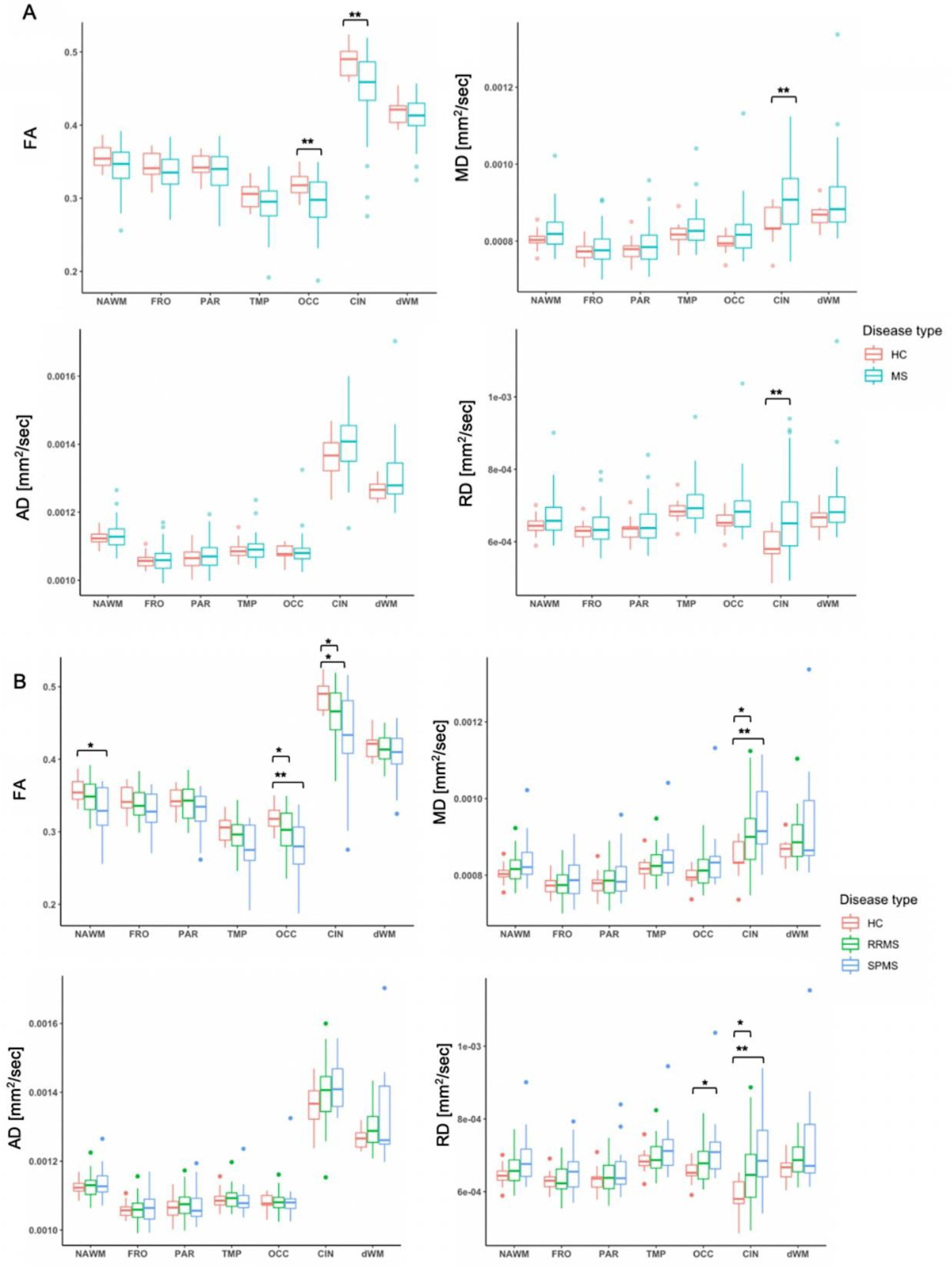
Structural changes in whole and segmented NAWM regions in MS patients compared to healthy controls. Differences in DTI parameters in the whole and segmented NAWM subregions of 55 MS patients compared to 15 healthy controls (A), and between RRMS (40), SPMS (15) and healthy controls (15) subgroups (B). Boxplots show median DTI values with first and third interquartile. Data beyond end of whiskers are outliers and plotted as points. The pairwise comparisons were performed with Wilcoxon’s test and were considered statistically significant at the level of *p*<0.05. Holm method was used to adjust the *p*-values for multiple comparisons. AD=axial diffusivity; CIN=cingulate; dWM=deep normal appearing white matter; FA=fractional anisotropy; FRO=frontal; MD=mean diffusivity; NAWM = normal appearing white matter; OCC=occipital; PAR=parietal; RD=radial diffusivity; TMP=temporal; *\*p*<0.05; ** *p*<0.01.

In the analyses of the segmented subregions of NAWM, FA was significantly decreased in occipital [mean (SD) 0.295 (0.033) vs. 0.319 (0.018); *p*=0.009] and cingulate [0.452 (0.050) vs. 0.487 (0.022); *p*=0.004] NAWM, and mean MD and RD were significantly increased [0.917 (0.091) vs. 0.847 (0.047); *p*=0.005 and 0.667 (0.103) vs 0.589 (0.045); *p*=0.004, respectively] in the cingulate NAWM of MS patients vs. HC (Figure 2A). In this and in the next paragraph mean MD and RD values are expressed in scientific notation of 10^−3^.

When the RRMS, SPMS and HC subgroups were compared to each other, we observed a significant difference in the whole NAWM, in occipital and cingulate WM areas, with statistical difference more pronounced in SPMS than in RRMS subgroups (Figure 2B). In detail, FA was significantly decreased in the whole NAWM of SPMS patients, compared to HC [mean (SD) 0.329 (0.034) vs. 0.356 (0.017); *p*=0.026], in the occipital NAWM [0.278 (0.039); *p*=0.005 and 0.302 (0.029); *p=*0.043 vs. 0.319 (0.018) for SPMS and RRMS, respectively], in cingulate NAWM of both SPMS and RRMS subgroups [0.426 (0.071); *p=*0.011 for SPMS and 0.462 (0.036); *p=*0.013 for RRMS vs. HC 0.487 (0.022)]. Mean MD was significantly increased in the cingulate NAWM of SPMS and RRMS patients compared to HC [0.947 (0.102); *p=*0.006 and 0.905 (0.085); *p=*0.020 vs. HC 0.847 (0.047), respectively]. RD was increased in the cingulate NAWM of both MS subgroups compared to controls [0.651 (0.086); *p=*0.015 for RRMS and 0.709 (0.127); *p=*0.007 for SPMS vs. HC 0.589 (0.045)] and in the occipital NAWM of the SPMS subgroup compared to HC [0.722 (0.098) vs. 0.655 (0.030); *p=*0.018].

### Decreased tract integrity and increased microglial activation in the NAWM is associated with higher disability in MS

Several significant correlations between the DTI parameters in the NAWM and disease severity were observed. Decreased mean FA in the NAWM correlated to higher EDSS (Spearman correlation ρ=-0.51, *p*<0.001), while increased diffusivities were also associated with higher EDSS (ρ=0.47, *p*<0.001 for MD; ρ=0.30, *p*=0.031 for AD; ρ=0.51, *p*<0.001 for RD) (Figure 3). Additionally, decreased FA correlated with higher MSSS (ρ=-0.32; *p*=0.018). Similarly, all diffusivities had positive correlations with MSSS: ρ=0.39, *p*=0.004 for MD; ρ=0.34, *p*=0.013 for AD; and ρ=0.38, *p=*0.005 for RD. In addition, increased [^11^C](R)-PK11195 DVR in the NAWM associated with higher EDSS (Spearman correlation ρ=0.37; *p*=0.005; Figure 3), but there was no significant correlation between global NAWM TSPO binding and MSSS (ρ=0.22; *p*=0.107) (Table e-3).

**Figure 3.**
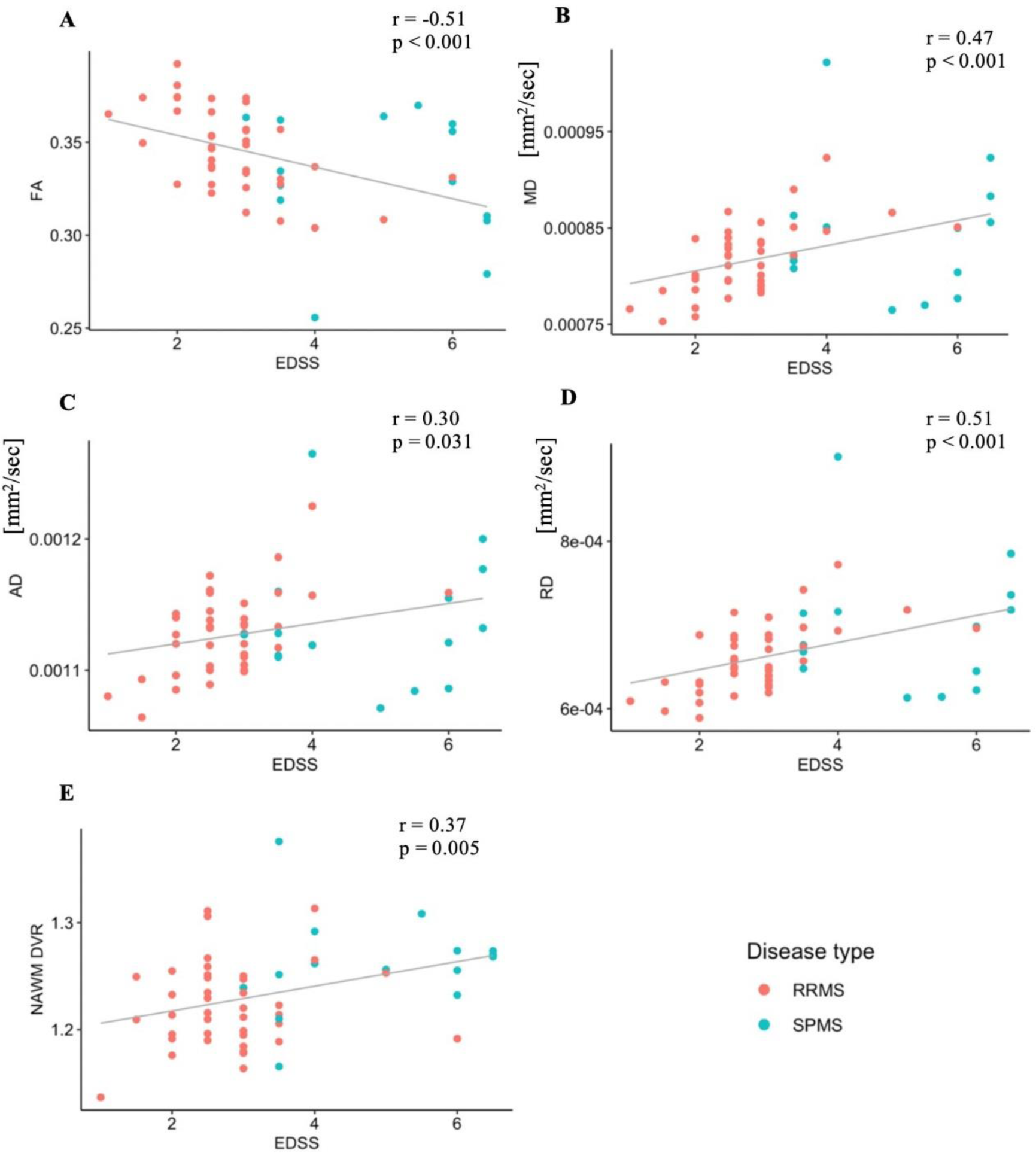
Associations of NAWM [^11^C](R)-PK11195 binding and decreased tract integrity to disability. Decreased WM integrity (A-D) and increased specific binding of [^11^C](R)-PK11195 measured as DVR in the global NAWM (E) is associated with clinical disability, evaluated with EDSS. The correlations are visualized with linear regression lines and Spearman correlation coefficient, significant at the level of *p*<0.05. AD=axial diffusivity; DVR=distribution volume ratio; EDSS=expanded disability status scale; FA=fractional anisotropy; MD=mean diffusivity; NAWM=normal appearing white matter; RD=radial diffusivity; RRMS=relapsing remitting multiple sclerosis; SPMS=secondary progressive multiple sclerosis.

Within the subregions of the NAWM, both decreased mean FA and increased diffusivity indices as well as increased [^11^C](R)-PK11195 DVR had several significant correlations with the disability and disease severity scores, the cingulate and occipital NAWM having the strongest associations (Table e-3).

### Increased microglial activation in the NAWM associates with structural white matter changes not visible in the conventional MRI in MS

The reduced WM tract integrity (demonstrated by reduced FA and increased MD, AD and RD) was significantly associated with increased [^11^C](R)-PK11195 DVR in the whole NAWM (ρ=-0.30, *p*=0.029 for FA; ρ=0.33, *p*=0.016 for MD; ρ=0.28, *p*=0.039 for AD; and ρ=0.34, *p*=0.013 for RD; Figure 4).

**Figure 4.**
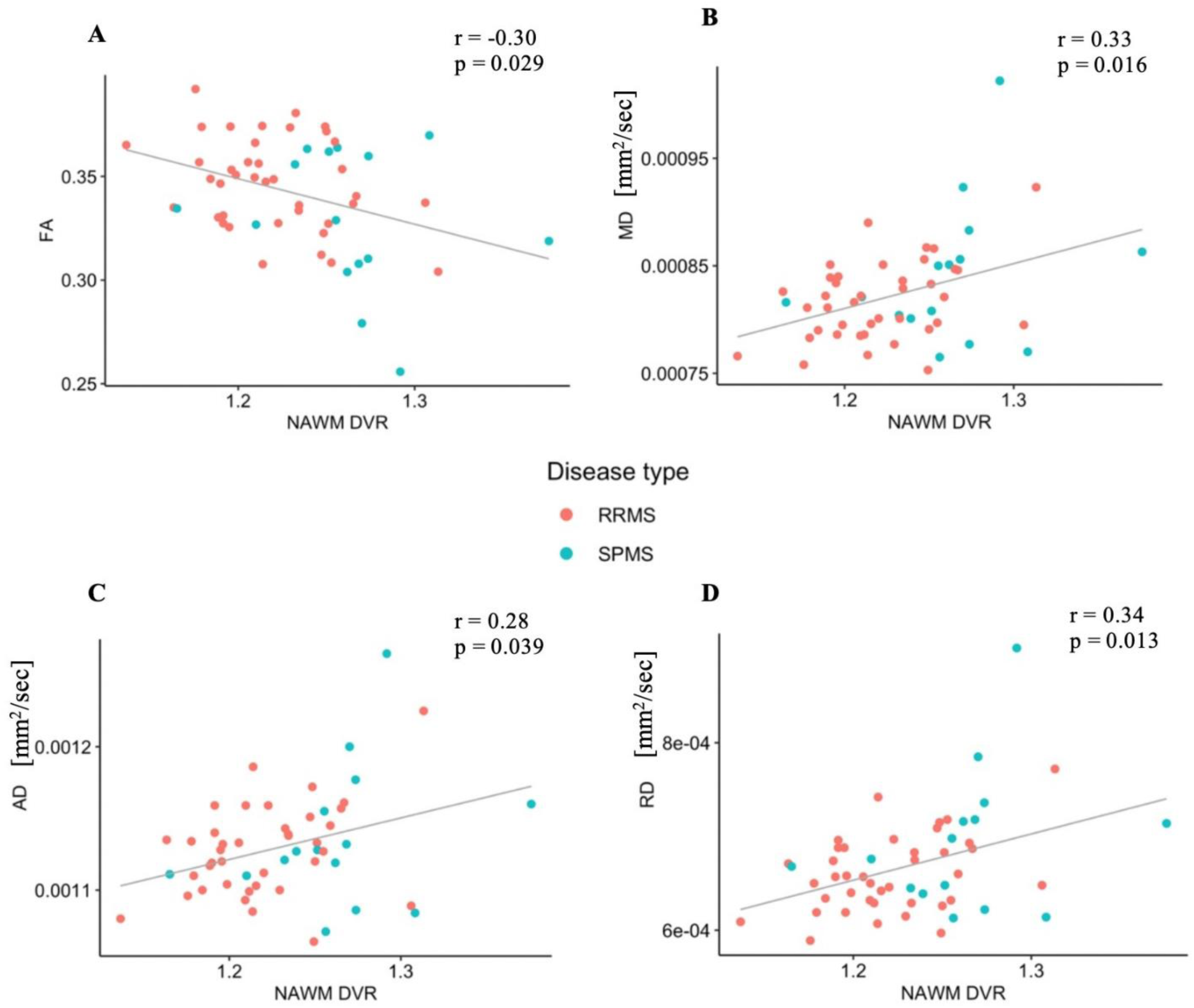
Associations between [^11^C](R)-PK11195 binding and structural white matter changes in NAWM in multiple sclerosis patients. Specific binding of [^11^C](R)-PK11195 measured as DVR in the NAWM correlates with fractional anisotropy (A), and with mean, axial and radial diffusivities (B-D) measured within the NAWM of 55 MS patients. The correlations are visualized with linear regression lines and Spearman correlation coefficient, significant at the level of *p*<0.05. AD=axial diffusivity; DVR=distribution volume ratio; FA=fractional anisotropy; MD=mean diffusivity; NAWM=normal appearing white matter; RD=radial diffusivity; RRMS=relapsing remitting multiple sclerosis; SPMS=secondary progressive multiple sclerosis.

Additionally, associations between increased [^11^C](R)-PK11195 DVR and decreased WM integrity were found in some NAWM subregions, most notably in the temporal NAWM (ρ=-0.39, *p*=0.02 for FA; ρ=0.40, *p*=0.014 for MD; ρ=0.37, *p*=0.036 for AD; ρ=0.43, *p*=0.007 for RD), data not shown.

### Voxel-wise image analysis of PET/DTI data

The voxel-level correlation analysis showed widespread focal areas of WM, where decreased FA correlated significantly with increased TSPO binding. Significant clusters were found within the superior longitudinal fasciculus, corticospinal tract, cingulate gyrus, corona radiata and forceps minor, and also within the thalamic radiation and the thalamus. The significant voxel-wise correlation coefficients are illustrated in Figure 5.

**Figure 5.**
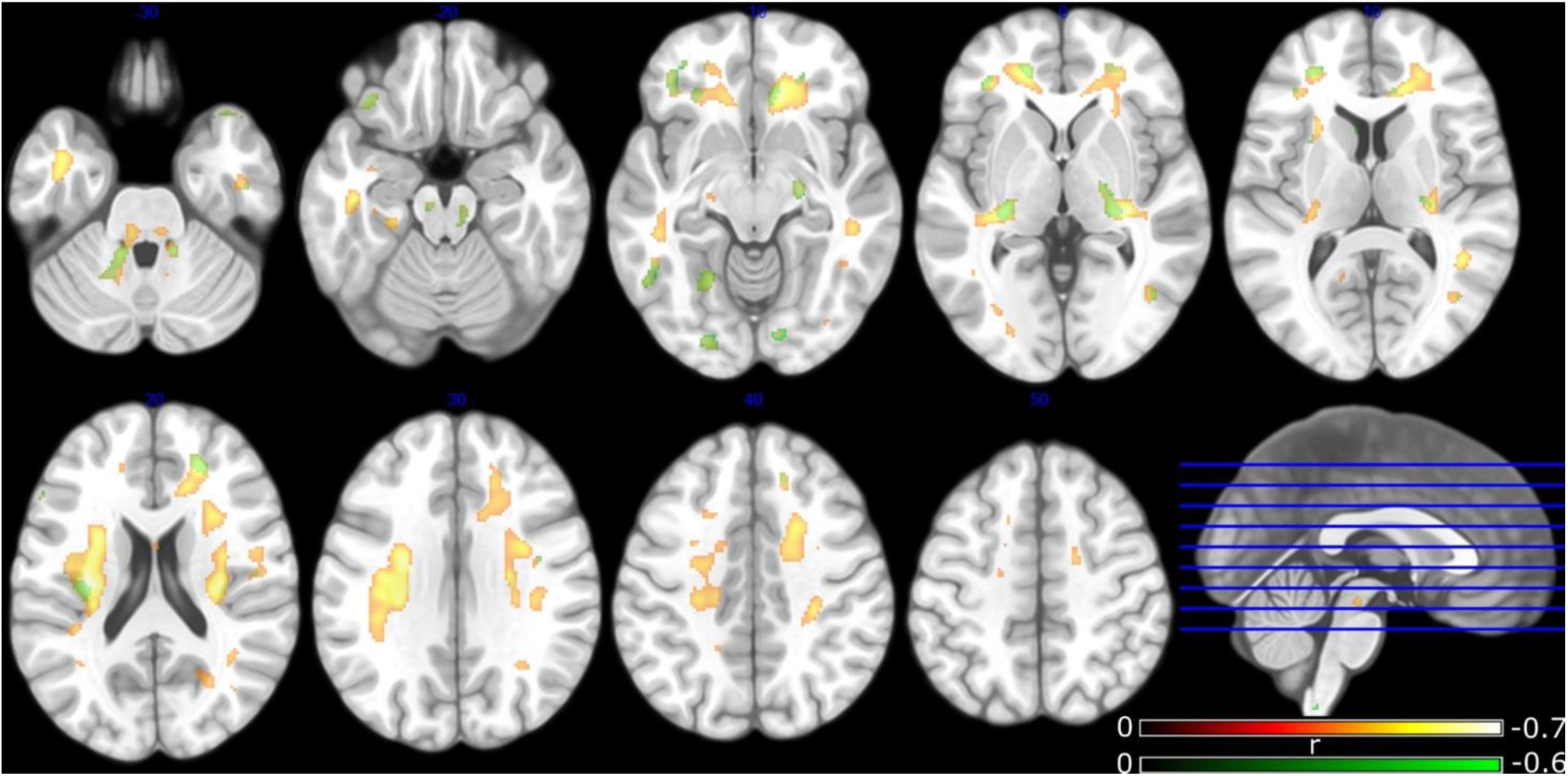
Voxel-wise correlation of increased microglial activation and decreased FA in the NAWM of MS patients. Statistically significant voxel-wise FA and DVR correlation coefficients in the NAWM (shown in green) and in the whole WM (shown in red-yellow) for the MS patients (n=54) at group level. Correction for multiple comparisons was performed using FDR with significance level of *p*<0.05. Significant clusters were found within the superior longitudinal fasciculus, corticospinal tract, cingulate gyrus, corona radiata and forceps minor, and also within thalamic radiation and thalamus. DVR=distribution volume ratio; FA=fractional anisotropy; FDR=false discovery rate; MS=multiple sclerosis; NAWM=normal appearing white matter; WM=white matter.

## DISCUSSION

This study evaluated structural and molecular brain changes using combined MR and PET *in vivo* imaging techniques in 55 MS patients having either RRMS (n=40) or SPMS (n=15). The results from the MS patients were compared to those from 15 healthy control subjects. Altered DTI parameters (decreased FA and increased MD, AD and RD) in the NAWM and in segmented subregions of the NAWM indicated microstructural damage without visible changes in cMRI, which is in agreement with previously published studies.^11,13,14^ Microstructural changes were more pronounced in SPMS compared to RRMS patients, and the most affected area was the cingulate NAWM region. The current understanding is that AD reports on axonal integrity, whereas RD changes associate with myelin integrity.^15^ Decreased FA has been interpreted to report on a decrease in axonal density, whereas an increase in MD associates with axonal and myelin loss.^15^ Our multimodal *in vivo* imaging findings hence contribute to the understanding of MS tissue pathology by demonstrating widespread damage both in myelin and axonal components, which associate with microglial activation within the NAWM. Our study also demonstrates, that the decreased WM tract integrity and microglial activation in the whole and in the segmented NAWM is associated with clinical disability and disease severity. Our work supports the present notion that CNS damage associated with MS disease progression is associated with chronic inflammation trapped within the CNS. The diffuse, widespread inflammation likely contributes to demyelination and axonal injury, as a consequence of chronic microglial activation, extensive production of reactive oxygen species (ROS) and iron accumulation.^16^ Inflammation is seen at all stages of the disease in chronic active MS lesions and in areas adjacent to them, i.e. also in the NAWM, with correlation between inflammation and axonal injury.^17^ For better understanding of the evolution of MS disease, it is important to be able to detect the diffuse neuroinflammation and associated axonal damage *in vivo* already prior to clinically evident progressive phase and prior to development of gross atrophy.

Some studies have demonstrated thalamic involvement in MS, with evidence of FA reduction in the thalamus^18,19^ that may predict disease evolution and contribute to cognitive decline and disability in MS. A common hypothesis is that focal lesions in thalamocortical projections can damage the surrounding WM, therefore leading to thalamic atrophy and neuronal loss.^20^ Results of the voxel-wise correlational analysis in this study demonstrated DTI pathology (reduced FA) in the thalami and the thalamic radiation in 54 MS patients, in association with increased TSPO binding as a measure of microglial activation. This is in line with the previous results on thalamic involvement in MS.^21^ In the voxel-wise correlation analysis the volume of voxels with statistically significant correlation between increased TSPO binding and decreased FA was larger in the whole white matter (including lesions) compared to the NAWM. It is most likely that the lower number of significant voxels in the NAWM compared to the whole WM is due to the largely varying location of lesions between each case, thus leaving less voxels for statistical comparison with the exact same brain co-ordinates belonging to NAWM in all cases. Consequently, this lower number of NAWM voxels with significant correlation reduces the statistical power and results in smaller significant voxel clusters. It is plausible that both inflammatory pathology and tract-related damage is greater in areas of acute focal demyelinating lesions, but a significant correlation between increased microglial activation and reduced FA in the NAWM areas surrounding the lesions could also be demonstrated. This likely indicates the presence of chronically active lesions with accumulation of activated microglia/macrophages^22^ and possibly Wallerian and retrograde degeneration in the vicinity of the lesions.^23^ Moreover, TSPO uptake is generally lower within the chronic WM lesions, especially in SPMS, than in the adjacent perilesional area and in more distant NAWM,^11^ but FA is also low due to demyelination. Thus, including the lesions into comparison should drive the correlation between TSPO binding and FA into the opposite direction than observed here (positive correlation instead of the observed negative correlation).

The transition of parenchymal microglia and/or infiltrating peripheral macrophages from their homeostatic (resting) state to the activated state is accompanied by the increased expression of 18kDa mitochondrial translocator protein, therefore making [C^11^](R)-PK11195 radioligand a highly specific molecular marker of microglia/macrophages activation, and consequently of neuroinflammation.^24^ The increased [C^11^](R)-PK11195 radioligand binding may also arise from denser microglial cellularity at sites of tissue damage.^25^ Moreover, TSPO has been demonstrated to be upregulated also on activated astrocytes. Therefore, the detected TSPO PET signal may originate from either cell type.^26^ Indeed, several animal studies^27,28^ have demonstrated upregulated TSPO expression in reactive astrocytes after nervous system injury, but at a significantly lower level and temporarily distinct, compared to microglia. The present understanding is that under pathological situations, activated microglia trigger astrocyte activation, and hence both can be simultaneously detected in MS pathology.^29^

To our knowledge, this is the first study evaluating diffuse structural and molecular changes in MS utilizing high magnetic field DTI and PET as complementary imaging modalities, performed on a large MS cohort (n=55) with comparison to healthy subjects. Combining different imaging modalities for the analysis of disease pathology in single individual patients has recently become an attractive approach in the neuroimaging field.^30,31^ This approach has great potential in the future for increasing our understanding of the evolution of the pathology associated with disease progression. Combination of complementary neuroimaging modalities provides a comprehensive examination of the brain structure and function *in vivo*. This study proves, that the application of DTI with TSPO-PET imaging for detecting hidden neuroinflammation reveals new insights into the complexity of MS pathology.

## CONCLUSIONS

This study provides evidence on the associations between DTI detectable abnormalities and microglial activation, measured with TSPO-PET *in vivo* in the NAWM of MS patients. The study demonstrates that the combination of PET and DTI imaging enables better appreciation of the hidden MS pathology not visible using conventional MRI. Future implementations of this study might be beneficial in revealing associations between fiber connectivity and enlargement of smouldering lesions with activated microglia. Combination of complementary techniques such as DTI and PET imaging may also be utilized as a tool for evaluation of disease progression in treatment trials of progressive MS.

## ACKNOWLEDGEMENTS

All the study participants and the expert staff at Turku PET Centre are gratefully acknowledged for making this study possible. We are also sincerely thankful to Professor Jarmo Hietala and Dr. Heikki Laurikainen for providing the additional healthy control DTI data for the analyses.

## REFERENCES

1. Thompson AJ, Baranzini SE, Geurts J, Hemmer B, Ciccarelli O. Multiple sclerosis. Lancet. Elsevier Ltd; 2018;391:1622–1636.

2. Filippi M, Rocca MA. Conventional MRI in multiple sclerosis. J Neuroimaging. 2007;17:3–9.

3. Le Bihan D, Mangin JF, Poupon C, et al. Diffusion tensor imaging: concepts and applications. J Magn Reson Imaging. 2001;13:534–546.

4. Ching ASC, Kuhnast B, Damont A, Roeda D, Tavitian B, Dollé F. Current paradigm of the 18-kDa translocator protein (TSPO) as a molecular target for PET imaging in neuroinflammation and neurodegenerative diseases. Insights Imaging. 2012;3:111–119.

5. Schmidt P, Gaser C, Arsic M, et al. An automated tool for detection of FLAIR-hyperintense white-matter lesions in Multiple Sclerosis. Neuroimage. Elsevier Inc.; 2012;59:3774–3783.

6. Salat DH, Greve DN, Pacheco JL, et al. Regional White Matter Volume Differences in nondemented aging and Alzheimer’s disease. Neuroimage. 2009;44:1247–1258.

7. Leemans A, Jones DK. The B-matrix must be rotated when correcting for subject motion in DTI data. Magn Reson Med. 2009;61:1336–1349.

8. Chang LC, Jones DK, Pierpaoli C. RESTORE: Robust estimation of tensors by outlier rejection. Magn Reson Med. 2005;53:1088–1095.

9. Chou M, Mori S. Effects of b-Value and Echo Time on Magnetic Resonance Diffusion Tensor Imaging-Derived Parameters at 1. 5 T : A Voxel-Wise Study. J Med Biol Eng. 2012;33:45–50.

10. Rissanen E, Tuisku J, Rokka J, et al. In Vivo Detection of Diffuse Inflammation in Secondary Progressive Multiple Sclerosis Using PET Imaging and the Radioligand 11C-PK11195. J Nucl Med. 2014;55:939–944..

11. Rissanen E, Tuisku J, Vahlberg T, et al. Microglial activation, white matter tract damage, and disability in MS. Neurol Neuroimmunol Neuroinflamm. 2018;5:1–10.

12. Gunn RN, Lammertsma AA, Cunningham VJ. Parametric Imaging of Ligand-Receptor Binding in PET Using a Simplified Reference Region Model. Neuroimage. 1997;6:279–287.

13. Roosendaal SD, Geurts JJG, Vrenken H, et al. Regional DTI differences in multiple sclerosis patients. Neuroimage. Elsevier Inc.; 2009;44:1397–1403.

14. Onu M, Roceanu A, Sboto-Frankenstein U, et al. Diffusion abnormality maps in demyelinating disease: Correlations with clinical scores. Eur J Radiol. Elsevier Ireland Ltd; 2012;81:e386–e391.

15. Sbardella E, Tona F, Petsas N, Pantano P. DTI Measurements in Multiple Sclerosis: Evaluation of Brain Damage and Clinical Implications. Mult Scler Int. 2013;2013:1–11.

16. Lassmann H. Multiple sclerosis pathology. Cold Spring Harb Perspect Med. 2018;8:1–16.

17. Frischer JM, Bramow S, Dal-Bianco A, et al. The relation between inflammation and neurodegeneration in multiple sclerosis brains. Brain. 2009;132:1175–1189.

18. Mesaros S, Rocca MA, Pagani E, et al. Thalamic damage predicts the evolution of primary-progressive multiple sclerosis at 5 years. Am J Neuroradiol. 2011;32:1016–1020.

19. Deppe M, Krämer J, Tenberge JG, et al. Early silent microstructural degeneration and atrophy of the thalamocortical network in multiple sclerosis. Hum Brain Mapp. 2016;37:1866–1879.

20. Minagar A, Barnett MH, Benedict RHB, et al. The thalamus and multiple sclerosis: Modern views on pathologic, imaging, and clinical aspects. Neurology. 2013;80:210–219.

21. Vercellino M, Masera S, Lorenzatti M, et al. Demyelination, inflammation, and neurodegeneration in multiple sclerosis deep gray matter. J Neuropathol Exp Neurol. 2009;68:489–502.

22. Zrzavy T, Hametner S, Wimmer I, Butovsky O, Weiner HL, Lassmann H. Loss of “homeostatic” microglia and patterns of their activation in active multiple sclerosis. Brain. 2017;140:1900–1913.

23. Singh S, Dallenga T, Winkler A, et al. Relationship of acute axonal damage, Wallerian degeneration, and clinical disability in multiple sclerosis. J Neuroinflammation. Journal of Neuroinflammation; 2017;14:1–15.

24. Liu GJ, Middleton RJ, Hatty CR, et al. The 18 kDa translocator protein, microglia and neuroinflammation. Brain Pathol. 2014;24:631–653.

25. Owen DR, Narayan N, Wells L, et al. Pro-inflammatory activation of primary microglia and macrophages increases 18 kDa translocator protein expression in rodents but not humans. J Cereb Blood Flow Metab. 2017;37:2679–2690.

26. Lavisse S, Guillermier M, Herard A-S, et al. Reactive Astrocytes Overexpress TSPO and Are Detected by TSPO Positron Emission Tomography Imaging. J Neurosci. 2012;32:10809–10818.

27. Maeda J, Higuchi M, Inaji M, et al. Phase-dependent roles of reactive microglia and astrocytes in nervous system injury as delineated by imaging of peripheral benzodiazepine receptor. Brain Res. 2007;1157:100–111.

28. Chen M-K, Guilarte TR. Imaging the Peripheral Benzodiazepine Receptor Response in Central Nervous System Demyelination and Remyelination. Toxicol Sci. 2006;91:532–539.

29. Peterson TC, Buckwalter MS, Panicker N, et al. Neurotoxic reactive astrocytes are induced by activated microglia. Nature. Nature Publishing Group; 2017;541:481–487.

30. Teipel S, Drzezga A, Grothe MJ, et al. Multimodal imaging in Alzheimer’s disease: Validity and usefulness for early detection. Lancet Neurol. 2015;14:1037–1053.

31. Uludağ K, Roebroeck A. General overview on the merits of multimodal neuroimaging data fusion. Neuroimage. 2014;102:3–10.

